# Time to commencement of effective treatment in patients with drug-resistant tuberculosis diagnosed in the Torres Strait / Papua New Guinea cross-border region

**DOI:** 10.1101/2022.01.16.22269389

**Authors:** J’Belle Foster, Diana Mendez, Ben J. Marais, Dunstan Peniyamina, Emma S. McBryde

## Abstract

**Introduction:** Delays between self-reported symptom onset and commencement of effective treatment contribute to ongoing tuberculosis (TB) transmission, which is a particular concern in patients with drug-resistant (DR)-TB. We assessed improvements in time to commencement of effective treatment in patients diagnosed with DR-TB in the Torres Strait / Papua New Guinea cross-border region.

**Methods:** All laboratory-confirmed DR-TB cases diagnosed in the Torres Strait between 1 March 2000 and 31 March 2020 were reviewed. We assessed the total time from self-reported onset of symptoms to effective treatment commencement in different programmatic time periods. Pairwise analyses and time to event proportional hazard calculations were used to explore the association between delays in median time to effective treatment, and selected variables. Data were further analysed to examine predictors of excessive treatment delay.

**Results:** The median number of days from self-reported onset of symptoms to effective treatment commencement was 124 days (interquartile range 51-214) over two decades. Between 2006 and 2012, most (57%) cases exceeded this ‘grand median’ while the median ‘time to treat’ in the most recent time period (2016-2020) was significantly reduced to 29 days (*p* <0.001). Although there was a reduction in the median ‘time to treat’ with the introduction of Xpert MTB/RIF (135 days pre-Xpert vs 67 days post-Xpert) this was not statistically significant (*p* 0.07). Establishment of the Torres and Cape TB Control Unit on Thursday Island (2016-2020) was significantly associated with reduced treatment delay, compared to previous TB program period (2000-2005 *p* <0.04; 2006-2012 *p* <0.001).

**Conclusion:** Minimising TB treatment delay in remote settings like the Torres Strait / Papua New Guinea cross-border region requires effective decentralised diagnosis and management structures. The results of this study suggest that the establishment of the Torres and Cape TB Control Unit on Thursday Island significantly improved time to commencement of effective TB treatment. Possible contributing factors include better TB education, cross-border communication and patient-centred care.

## Introduction

Tuberculosis (TB) remains a disease of public health significance, with an estimated 10 million cases diagnosed in 2019 [1]. Australia’s TB incidence is considered low at 5.5/100,000 population [2] however, TB disproportionately affects Indigenous Australian populations, and the incidence rate in the Torres Strait in 2014 was 107/100,000 population (based on 2011 Census population data). In Papua New Guinea (PNG), TB is the leading cause of death from an infectious agent, with a TB case notification rate of 674/100,000 population in the Western Province in 2016 [3]. In 2014, the PNG National Department of Health declared Daru Island in the Western Province a ‘hot-spot’ as, of approximately 500 cases of TB diagnosed on the island each year, one in ten TB cases were multidrug-resistant (MDR) [4].

Disease control is impeded when drug-resistant (DR)-TB is poorly managed. DR-TB can be community-acquired, or be the result of irregular or interrupted treatment, inadequate drug regimens or malabsorption of treatment [5, 6]. Most types of DR-TB, including rifampicin resistant (RR) and multidrug-resistant TB (MDR-TB), require second-line drugs to treat which are more expensive, toxic, and the length of treatment is time-consuming [7]. MDR-TB is TB that is resistant to both isoniazid and rifampicin which are two of the most potent TB drugs [8]. Two major factors of treatment success for DR-TB patients are prompt diagnosis and commencement of effective sustained treatment [6]. Delays in treatment commencement can result in high bacillary load with increased infectiousness and spread [9]. Early effective treatment reduces the bacterial burden and limits transmission [8].

The Australian National TB Advisory Committee considers time to effective treatment commencement to be one of the most important markers of effective TB control programs [11]. In settings with excessive delays from symptom onset to treatment commencement, higher drug-resistance is expected [10]. Currently, there is no global consensus on how to define excessive delay from self-reported symptom onset to treatment commencement. In principle it is important to minimise treatment delay to limit transmission risk [12, 13]. Patient factors such as reduced health-seeking behaviours due to poor knowledge, distrust or other impediments may delay presentation [13, 14]. Health-system related factors such as poor TB awareness by staff, or a lack of appropriate laboratory facilities, as well as radiology and treatment access, can also contribute to treatment delay [14, 15]. Individual and health-system related factors that lead to delays should be monitored and addressed by TB control units [11].

Geographical and geopolitical challenges in the Torres Strait / PNG region create health care disparities which affect diagnosis, referral pathways, treatment and management of TB patients in the Torres Strait Protected Zone (TSPZ). Since the ratification of the Torres Strait Treaty in 1985, free bidirectional movement has been allowed for traditional, family, economic and trade purposes in the TSPZ between 13 PNG Treaty villages in the Western Province and 13 adjacent Australian Torres Strait Islands [16, 17]. As a result of this international agreement, TB has often been able to move, unhindered, across borders [18]. Primary transmission of TB and the rise of drug-resistance are cause for concern for at-risk Australian and PNG border communities [19]. The Torres Strait Treaty does not include access to Australian healthcare for PNG villagers living adjacent to the Australian border, despite the narrow (4.7 kilometre) stretch of water separating the two countries (refer to figure 1). In response to the urgent nature of some healthcare needs, Queensland Health provides humanitarian aid to those most in need as well as providing triaging and point of care diagnostic services.

**Figure 1.**
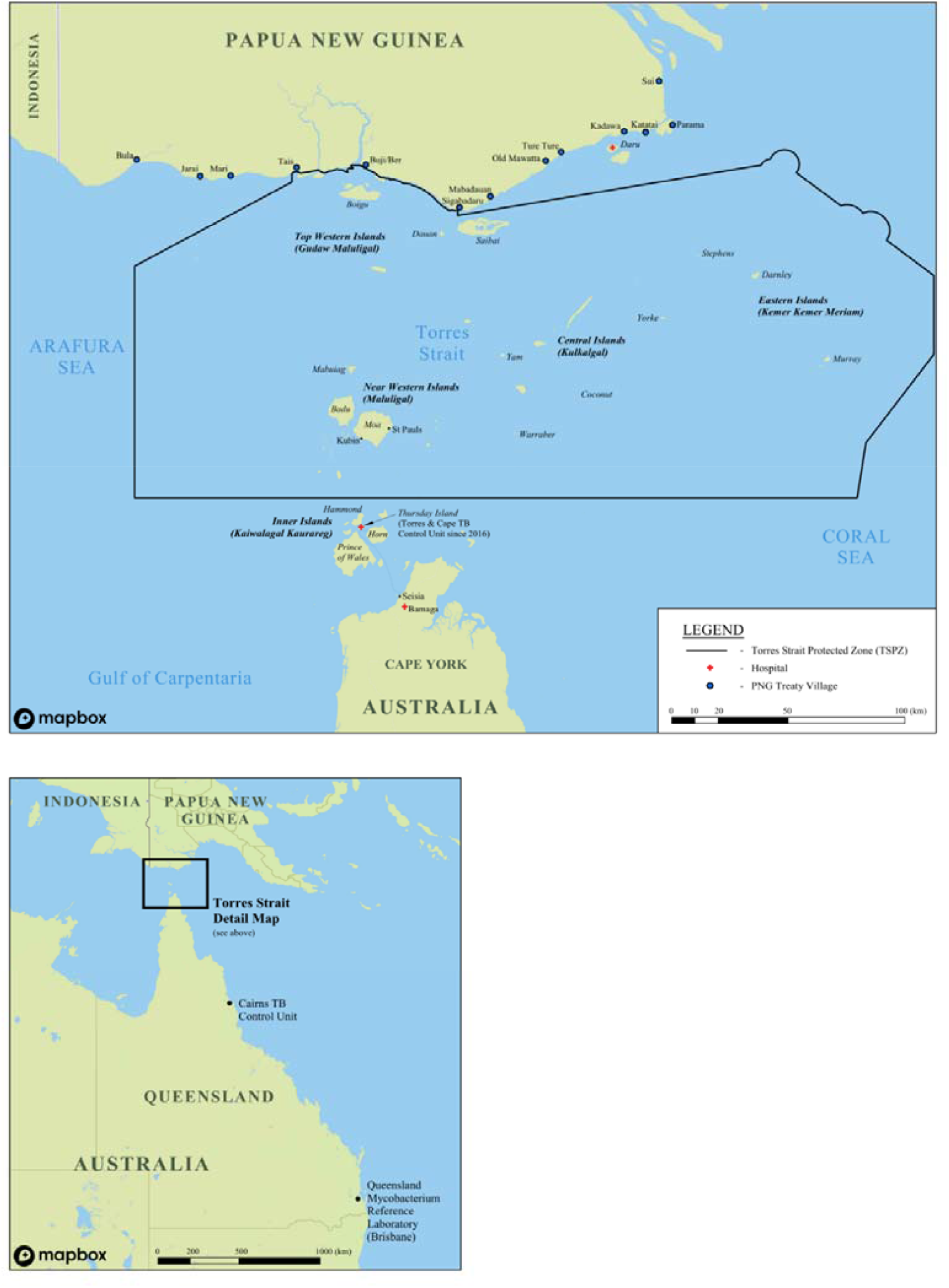
Map of the Torres Strait / Papua New Guinea cross-border region^1^. ^1^ DOI. 10.6084/m9.figshare.16632823

Optimising models of care for TB patients living in the Torres Strait / PNG international border region has been challenging and the programmatic TB response in the remote area has evolved over time (refer to figure 2). Although Torres Strait and PNG cross-border communities are subject to a similar TB risk they do not have access to the same level of health care [20]. On the Australian side of the border, high-risk screening and passive TB case finding is standard practice whereby cases are diagnosed post-investigation of TB-related symptoms or incidentally while other medical conditions are being investigated [11, 21]. During the study period, PNG nationals requiring TB work-up who did not present to an Australian health facility needed to travel to Daru General Hospital, located at least two hours away by small motorised boat (at a cost of AUD$240) [22]. The region’s geography and associated travel costs renders accessing TB health care services prohibitive for PNG nationals residing adjacent to the TSPZ. The Multi-dimensional Poverty Index in the area is estimated to be 0.32 [23]. Over the past 20 years, poverty has affected time to treatment commencement for PNG residents with TB living adjacent to the Australian border. The consequences are sustained transmission risk and rapid growth of DR-TB on both sides of the border.

**Figure 2.**
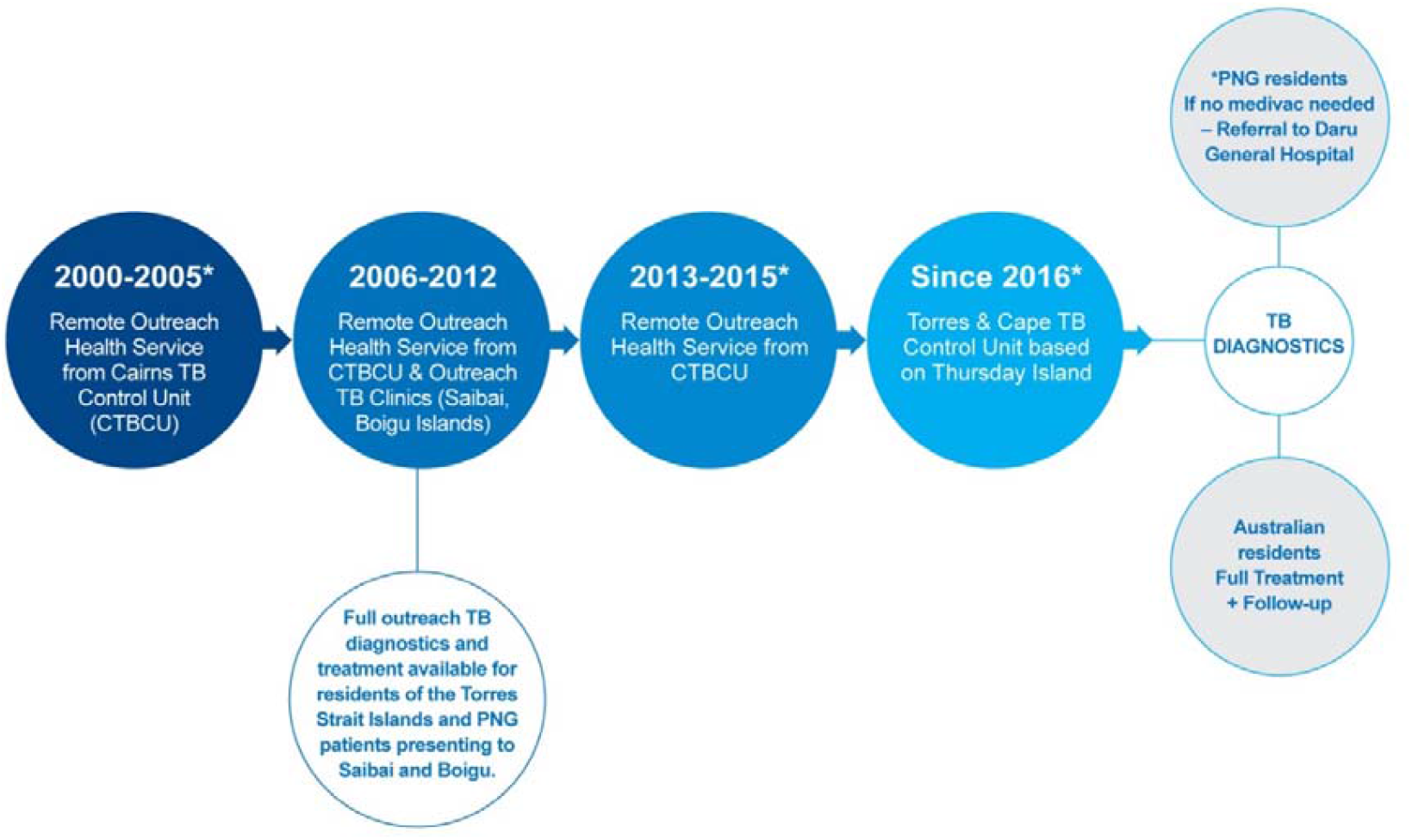
Description of the different tuberculosis programmatic time periods in the Torres Strait / Papua New Guinea cross-border region between 2000 and 2020^2^. ^2^ DOI. 10.6084/m9.figshare.16834648

Time from self-reported onset of symptoms to effective treatment commencement has not previously been investigated in this particular context. This study aimed to explore total time from self-reported onset of TB symptoms to effective treatment commencement for all DR-TB cases diagnosed in the Torres Strait / PNG border region over two decades. This study identifies factors associated with the time between self-reported symptom onset to effective treatment commencement as well as discusses programmatic interventions that may have contributed to reduced treatment delays over time. As DR-TB transmission is an established risk in the Torres Strait / PNG border, this study may help to improve programmatic management of TB in the region to the benefit of both Australian and PNG health care systems and local communities.

## Methods

### Design and Study Population

This retrospective cohort study included all patients from Australia or PNG that were diagnosed in the Torres Strait Islands with microbiologically-confirmed DR-TB between 1 March 2000 and 31 March 2020. Patients of all ages, including children, were included in the study, as were those with pulmonary, extrapulmonary, smear negative and smear positive disease.

Sputum and other histological samples were collected in the Torres Strait Islands and transported via sea and air to a WHO-designated Supranational Reference Laboratory, the Queensland Mycobacterium Reference Laboratory (QMRL) [24] (refer to figure 1). Confirmation of DR-TB diagnosis was confirmed by culture and phenotypic drug susceptibility testing (DST) pre-November 2010, or by Cepheid Xpert® MTB/Rif Assay (Xpert) (rapid diagnostic technology), culture and DST post-November 2010 [24].

### Definitions

The sample size is based on 133 cases with DR-TB, with 113 experiencing the event (known effective treatment commencement).

*Effective treatment* was defined as the implementation of appropriate second-line TB treatment for RR/MDR-TB cases and appropriate TB treatment for other drug-resistant cases, whether this was programmatic / empiric or personalised.

*Time to event* was defined as the sum in days of the following three distinct intervals:

1. Patient Delay (the time from self-reported TB symptom onset to presentation at a health facility in the Torres Strait)
2. Health System Delay (the time from presentation at a health facility to being diagnosed with TB)
3. Treatment Delay (the time from diagnosis to event – effective treatment commencement)

As not all participants commenced treatment, the data are right censored. The 20 censored cases that were transferred out or died were observed in the study to pass through the sum (in days) of patient delay and health system delay – time from self-reported onset of symptoms to diagnosis.

*Diagnosis year group* reflects the changes in programmatic TB responses in the remote Torres Strait / PNG international border that occurred over time (refer to figure 2).

*Case type* was defined as either a new case, or relapsed case following full or partial treatment in Australia or overseas. These were analysed as independent events. Four patients who had previously received full or partial treatment were diagnosed twice in this study, and were counted discretely as separate events. Self-reported onset of symptoms did not overlap for any of these four patients.

### Data Collection

Clinical and demographic data were sourced from the Queensland Notifiable Conditions System (NoCS), and drug-resistance for each case was verified in Queensland Health’s laboratory results software AUSLAB. Case notifications were cross-checked between NoCS and AUSLAB and one case of DR-TB identified from NoCS but not from AUSLAB, was removed from the study.

The date of symptom onset and treatment commencement in NoCS was cross-checked against self-reported symptom onset date and treatment commencement documented in patient charts, Queensland Health’s state-wide patient database, The Viewer [25], and in the Torres and Cape Hospital and Health Service (North) patient database, Best Practice [26]. As NoCS relied on retrospective submissions of case notification data throughout the study period, original or copies of handwritten progress notes in patient charts, and referrals and progress reports in The Viewer and Best Practice were considered the sources of truth for this study for symptom onset and treatment commencement. If this evidence was not available, the date of symptom onset and treatment commencement in NoCS was used.

For data pertaining to 2016 and onwards, a combination of NoCS, The Viewer, Best Practice and two Excel spreadsheets specifically used to manage patient data for residents of the Torres Strait Islands, and PNG patients diagnosed with TB, were used to ascertain self-reported date of onset of symptoms and date of treatment commencement for each case. Prior to the border closures associated with the COVID-19 pandemic, clinicians from the Torres and Cape TB Control Unit met with clinicians from the Daru TB Programme to discuss treatment commencement dates and treatment outcomes of shared cross-border patients. Further, a core component of the Torres and Cape TB Control Unit’s cross-border TB portfolio, was to share new laboratory results, chest x-ray reports and household contact information with the Daru TB Programme.

Patients diagnosed with DR-TB at an Australian health facility prior to the end of 2012, commenced treatment for DR-TB in Australia as access to second-line drugs used to treat DR-TB were not yet available on Daru Island. Once drug-susceptibility testing and effective treatment of DR-TB were available on Daru Island, the numbers of DR-TB cases diagnosed by Australian health facilities decreased. Most patients from PNG who were diagnosed at Australian health facilities with DR-TB between 2013 and 2020 commenced treatment in PNG, however, the most critically unwell that were transferred to either Thursday Island, Cairns or Townsville Hospitals may have commenced treatment in Australia. We did not collect data pertaining to which site the patient commenced effective treatment.

## Data Analyses

All statistical analyses were performed using IBM SPSS Statistics, version 27 (2020, Armonk, New York, United States).

Frequencies and percentages were calculated for categorical variables including treatment event (effective treatment commencement or transfer out / death), visa status, case type, RR-TB, MDR-TB and gender.

Tests for normality were performed for mean, median, skewness and kurtosis, and visual inspections of histograms were reviewed. Assumptions for normality were violated and data were not normally distributed. Medians and interquartile range (IQR) were used for data that were not normally distributed to demonstrate differences in the total time to effective treatment commencement across the four different time periods. When determining median days to effective treatment commencement, patients who did not commence known treatment and were transferred out or died, were removed from analyses, hence time to treatment outcomes should be interpreted as conditional on treatment commencement.

To determine any association between advanced diagnostic technology implementation in Queensland from November 2010, a non-parametric independent samples Median Test for k samples was used. The dependent variable was total time in days to known treatment commencement and the independent categorical variables were pre and post-Xpert. Patients that did not commence effective TB treatment due to death or transfer out were removed from the analyses.

Independent samples median test of frequencies was used to both identify the grand median, and to demonstrate frequencies above and below the median days to effective treatment commencement across different time periods. Post-hoc analyses were performed using pairwise comparison to determine the differences of median delays across the four different time periods, reflecting different programmatic management types. The dependent variable was total time to event (symptom onset to effective treatment commencement), and two time periods were compared at a time using pairwise comparison. Statistical significance was set at *p* <0.05. Due to the relatively small sample size and to counter a potential Type 1 error that may occur when multiple analyses are performed on the same dependent variable, a Bonferri correction was applied [27]. The alpha was lowered by taking the unadjusted p-value and multiplying it by the number of pairwise comparisons [27].

A Cox proportional hazards regression plot was used to illustrate the time to known treatment commencement in days, stratified by case type (new versus previous full or partial treatment).

## Ethics

This study was conducted with the ethical approval from the Far North Queensland Human Research Ethics Committee (HREC) (HREC/17/QCH/74-1157), the Chair of James Cook University HREC (H7380) and a Public Health Act approval (QCH/36155 – 1157).

## Results

### Descriptive Statistics

Table 1 shows that of 133 patients diagnosed with DR-TB between 2000 and 2020, 20% had previously received full or partial TB treatment in Australia or overseas and 70% were resistant to rifampicin. The proportion of patients with MDR-TB overall who previously received full or partial treatment was 26% (23/89 cases). Between 2000 and 2005, 100% of DR-TB cases were new and of those, 71% were RR.

**Table 1.**
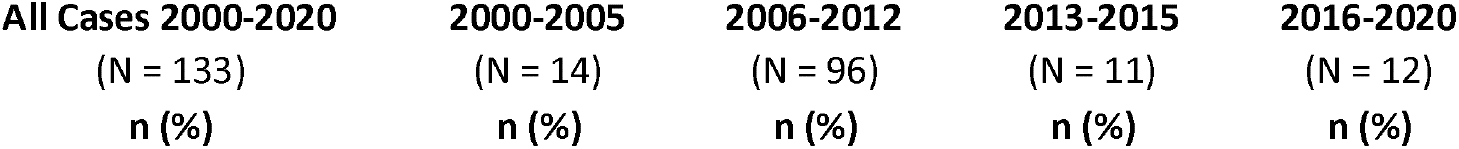
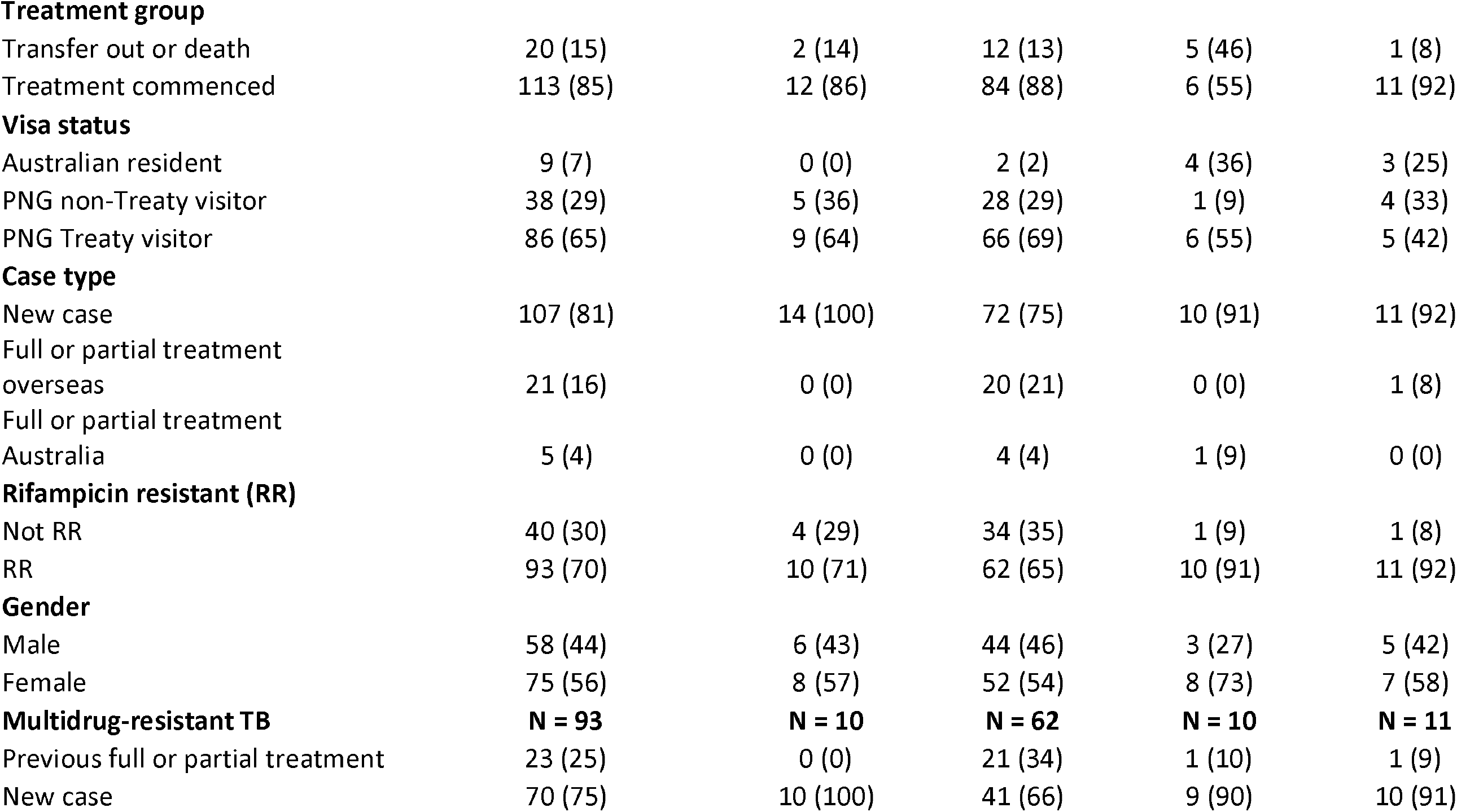
Characteristics of drug-resistant tuberculosis patients diagnosed during different programmatic time period in the Torres Strait / Papua New Guinea cross-border region between 2000 and 2020.

| **All Cases 2000-2020** | **2000-2005** | **2006-2012** | **2013-2015** | **2016-2020** |
| --- | --- | --- | --- | --- |
| (N = 133) | (N = 14) | (N = 96) | (N = 11) | (N = 12) |
| n (%) | n (%) | n (%) | n (%) | n (%) |

|  |  |  |  |  |  |
| --- | --- | --- | --- | --- | --- |
| **Treatment group** |  |  |  |  |  |
| Transfer out or death | 20 (15) | 2 (14) | 12 (13) | 5 (46) | 1 (8) |
| Treatment commenced | 113 (85) | 12 (86) | 84 (88) | 6 (55) | 11 (92) |
| **Visa status** |  |  |  |  |  |
| Australian resident | 9 (7) | 0 (0) | 2 (2) | 4 (36) | 3 (25) |
| PNG non-Treaty visitor | 38 (29) | 5 (36) | 28 (29) | 1 (9) | 4 (33) |
| PNG Treaty visitor | 86 (65) | 9 (64) | 66 (69) | 6 (55) | 5 (42) |
| **Case type** |  |  |  |  |  |
| New case | 107 (81) | 14 (100) | 72 (75) | 10 (91) | 11 (92) |
| Full or partial treatment overseas | 21 (16) | 0 (0) | 20 (21) | 0 (0) | 1 (8) |
| Full or partial treatment Australia | 5 (4) | 0 (0) | 4 (4) | 1 (9) | 0 (0) |
| **Rifampicin resistant (RR)** |  |  |  |  |  |
| Not RR | 40 (30) | 4 (29) | 34 (35) | 1 (9) | 1 (8) |
| RR | 93 (70) | 10 (71) | 62 (65) | 10 (91) | 11 (92) |
| **Gender** |  |  |  |  |  |
| Male | 58 (44) | 6 (43) | 44 (46) | 3 (27) | 5 (42) |
| Female | 75 (56) | 8 (57) | 52 (54) | 8 (73) | 7 (58) |
| **Multidrug-resistant TB** | **N = 93** | **N = 10** | **N = 62** | **N = 10** | **N = 11** |
| Previous full or partial treatment | 23 (25) | 0 (0) | 21 (34) | 1 (10) | 1 (9) |
| New case | 70 (75) | 10 (100) | 41 (66) | 9 (90) | 10 (91) |

Table 2 demonstrates that the median time to effective treatment commencement for 113 patients increased after 2000 to 2005 (92 days; IQR 70-201) and peaked between 2006 and 2012 at 138 days (IQR 72-251). Reduction to 29 days (IQR 7-45) was observed between 2016 and 2020.

**Table 2.** Time to commencement of effective drug-resistant tuberculosis treatment across different tuberculosis programmatic time periods in the Torres Strait / Papua New Guinea cross-border region between 2000 and 2020.

| Variable | Time period* |  |  |  |
| --- | --- | --- | --- | --- |
|  | Services coordinated from Cairns |  |  | Thursday Island coordination |
|  | 2000-2005 | 2006-2012 | 2013-2015 | 2016-2020 |
| **N** | 12 | 84 | 6 | 11 |
| **Median (IQR)** | 92 (70-201) | 138 (72-251) | 67 (46-198) | 29 (7-45) |

\*See figure 2 for description of tuberculosis programmatic time periods in the Torres Strait / Papua New Guinea cross-border region between 2000 and 2020

### Inferential Statistics

The independent samples median test (figure 3) shows that the grand median time from self-reported TB symptom onset to known effective treatment commencement for DR-TB cases was 124 days (IQR 51-214). Figure 3 shows post-hoc pairwise analyses demonstrating a statistically significant difference between the “time to known effective treatment commencement” medians of patients in the diagnosis year groups 2000-2005 and 2016-2020 (*p* 0.04), and year groups 2006-2012 and 2016-2020 (*p* <0.001). Frequencies of cases both above and below the median are illustrated in figure 3. Fifty-seven percent (n = 48) of cases diagnosed between 2006 and 2012 exceeded the median total time to effective treatment commencement. All other diagnosis year groups had greater numbers of participants below the median than above the median.

**Figure 3.**
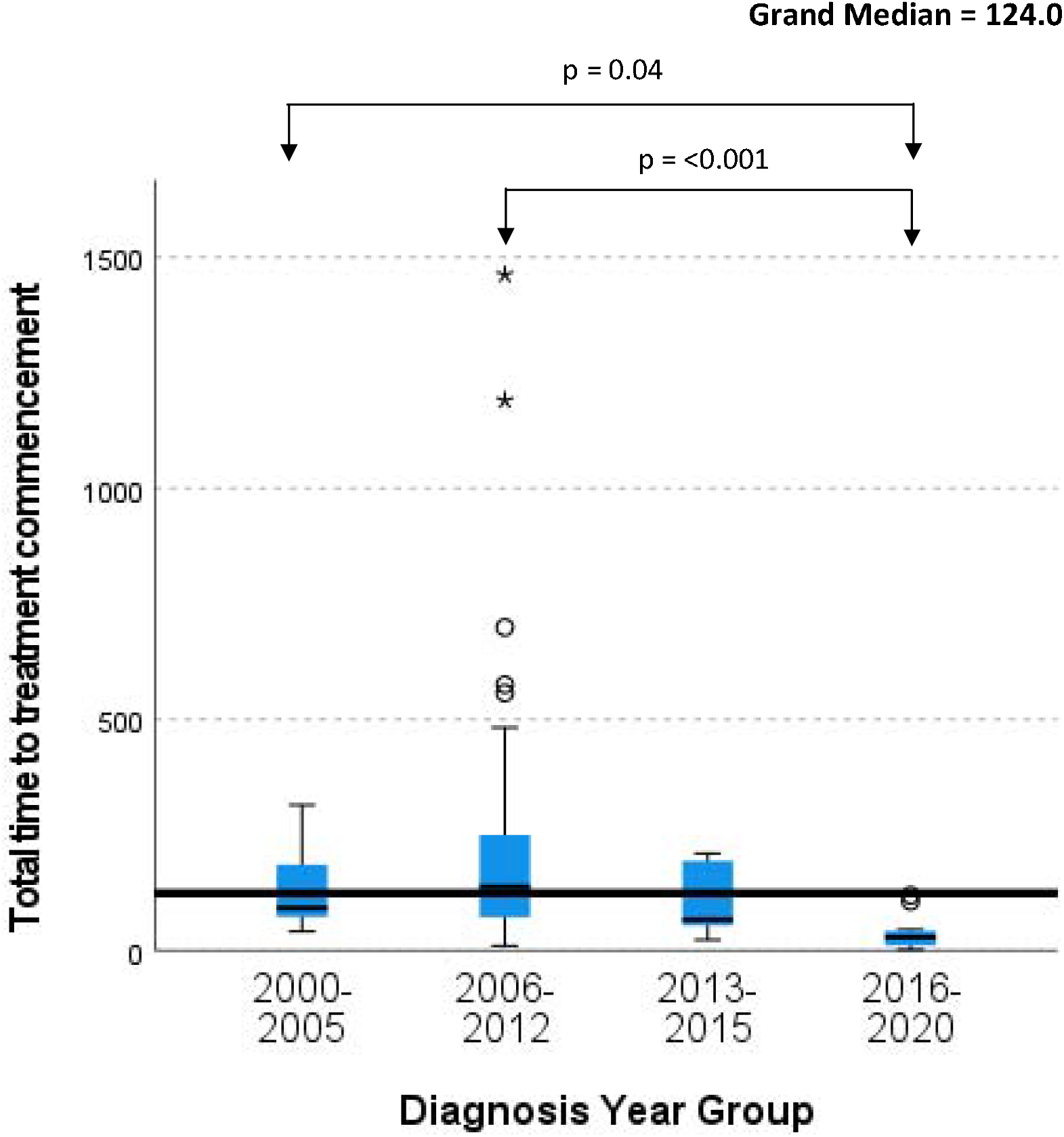
Variability in median time to treatment commencement across different tuberculosis programmatic time periods in the Torres Strait / Papua New Guinea cross border region between 2000 and 2020. Grand median – the pooled median in days of all drug-resistant TB cases that commenced effective tuberculosis treatment Significance values have been adjusted by the Bonferroni correction for the correct level of significance for multiple tests. Non-significant results are not shown.

The independent samples median test (table 3) demonstrates that post-implementation of the rapid diagnostic technology, Xpert, the median time from self-reported symptom onset to treatment commencement was 67 days (IQR 30-202) and the pre-Xpert time to treat median was 135 days (IQR 73-248; *p* 0.07).

**Table 3.** Time to commencement of effective drug-resistant tuberculosis treatment pre and post-Xpert availability in the Torres Strait / Papua New Guinea cross-border region between 2000 and 2020.

|  | **Pre-Xpert (2000-Oct 2010)** | **Post-Xpert (Nov 2010 – 2020)** | **P-value** |
| --- | --- | --- | --- |
| **N** | 75 | 38 |  |
| **Median (IQR)** | 135 (73-248) | 67 (30-202) | 0.07 |

Note. Xpert® MTB/RIF assay was performed in Cairns or Brisbane

Figure 4a shows the Cox proportional hazard plot for case type and total time to commencement of effective treatment. As can be seen in this figure, patients that had previously received full or partial treatment, demonstrated a more gradual slope to hazard (i.e., “time to known commencement of effective treatment”). Case type emerged as a predictor of earlier effective treatment commencement (HR = 0.6 (95%CI 0.35-0.89); p 0.01). Figure 4b shows the Cox proportional hazard plot for TB services provided by Cairns TB Control Unit and the Torres and Cape TB Control Unit. This figure shows that patients treated within the Cairns TB Control Unit, demonstrated a more gradual slope to hazard (i.e. “time to known commencement of effective treatment”).

**Figure 4a.**
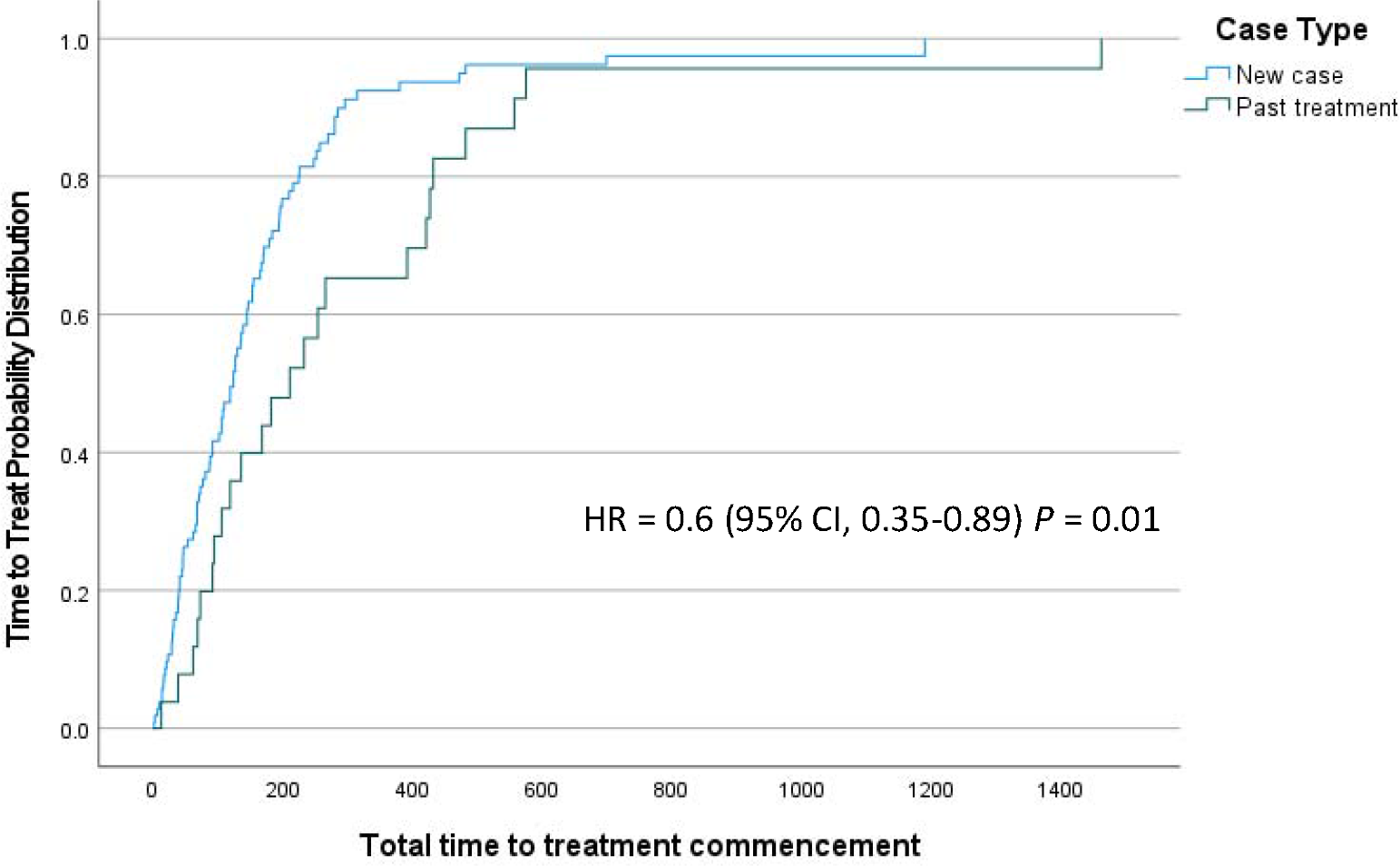
Time to treat (days) stratified by ‘new case’ and ‘past treatment’ in the Torres Strait / Papua New Guinea cross border region between 2000 and 2020. New case – without any previous TB treatment Past treatment – received full or partial TB treatment in the past

**Figure 4b.**
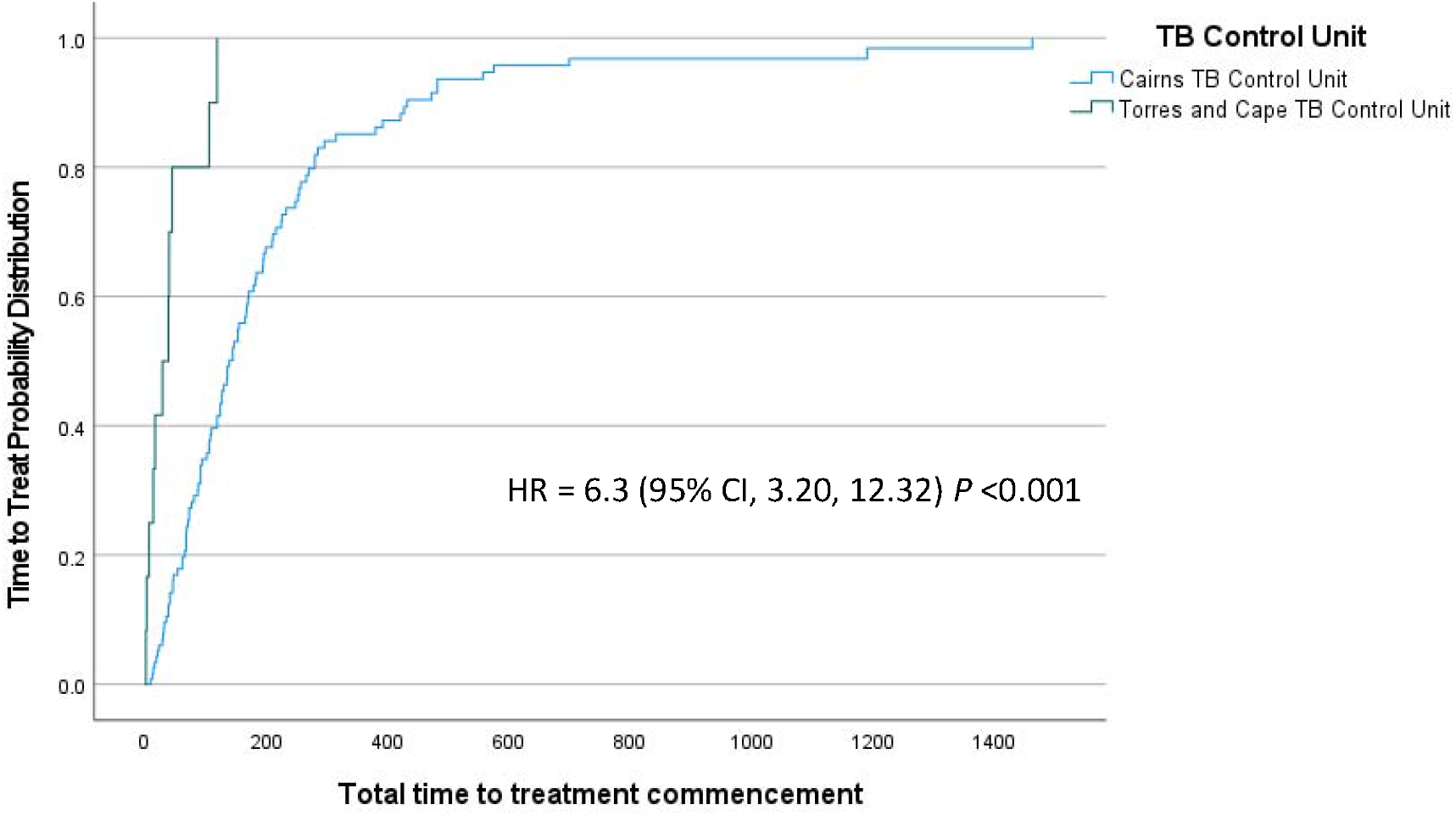
Time to treat (days) stratified by TB services provided by the ‘Cairns Tuberculosis Control Unit and Torres and Cape Tuberculosis Control Unit’ in the Torres Strait / Papua New Guinea cross border region between 2000 and 2020.

## Discussion

In this study, the lengthy median delay from self-reported onset of symptoms to effective treatment commencement was 124 days (IQR 51-214) over the study period. This is approximately four times more than the time to treat in a study conducted in the late 1990’s in metropolitan Victoria, Australia, where 33 days from symptom onset to treatment commencement was deemed an acceptable delay [28]. Even considering remoteness this key finding is still well above delays in other similar settings, and we offer 4-6 weeks as an acceptable delay, based on results from Australian literature. A systematic review of 198 studies reported a pooled mean total delay of 88 days across low and middle-income countries in both urban and remote settings [12]. Remoteness of the region, diagnostic services located on mainland Australia, access to pharmaceuticals for patients who commenced treatment in Australia and limited telecommunications with residents of Treaty and non-Treaty villages in PNG all remained unchanged throughout the study period, therefore variation of time to effective treatment are likely to be associated with other factors. These may be low-level of health service coverage and lack of awareness of TB as has previously been identified in other rural and remote settings [14].

Excessive delays between onset of symptoms and effective treatment commencement may contribute to ongoing transmission of TB in the region [15], hence various programmatic initiatives were introduced by the Torres and Cape TB Control Unit to strive to reduce this time. For example, culturally competent health promotion has been shown to increase knowledge and uptake of disease prevention strategies in culturally diverse communities [29]. Initiatives introduced by the Torres and Cape TB Control Unit included specific onboarding and opportunistic education for staff, and development and distribution of linguistically and culturally appropriate TB education materials. These initiatives may have been contributing factors to the decrease in time to effective treatment (29 days) from 2016 to below the level considered acceptable in metropolitan Victoria (33 days) [28]. These contributing factors may require further investigation to better understand their effect on time to treatment.

Changes to the management of TB in the region over time may help to explain a near 5-fold decrease in the median time to treat between 2006 to 2012 and 2016 to 2020. By 2012, the Daru TB Program in PNG, supported by the Australian Government, had developed capacity to manage both fully-susceptible and DR-TB cases [22]. Measures introduced in the region from 2014 and programmatic changes from 2016 may have also contributed to a reduction in median time from onset of symptoms to effective treatment commencement. From 2014-2016, Australian Commonwealth funding supported the delivery of TB-clinician-led initiatives to enhance TB knowledge amongst local communities and healthcare facilities on both sides of the international border [20, 30-32]. This enhanced knowledge of the basic features of TB may have assisted in reducing the time from self-reported onset of symptoms to presentation at a health facility.

During this time, Australian Commonwealth funding also supported a data integrity project [33] to gather all historic PNG patient charts located on Australian Torres Strait Islands and patient data contained in an electronic medical record used prior to 2011, Holt’s database [26]. Data migration into Best Practice software enabled clinicians to access patient information in a single electronic database. As a result of the cross-border data integrity project, from 2015, all PNG Nationals presenting with signs and symptoms of TB to a PHC in the TSPZ have an electronic medical record. This initiative led to increased real-time visibility of all patients in the TB care pathway, provided access to information about past TB diagnoses and past contact history dating back to the 1950’s, and allowed for identification and follow-up of previously treated cases.

Notably, the absence of advanced diagnostic technology such as Cepheid Xpert® MTB/Rif Assay that can identify RR in less than two hours, can add considerable time to health system related delays [34]. Implementation of Xpert in Queensland, Australia in November 2010 improved the overall median delay to 67 days in this study, which was approximately half the median pre-Xpert, indicating that implementation of Xpert in the Torres Strait may have helped to further reduce time to effective treatment delays [35]. Although the point-estimate difference pre and post-Xpert was large, there were additional temporal trends at the time and the difference pre- and post-Xpert was not statistically significant in this study.

Outreach TB specialist services provided by staff based in Cairns prior to 2016 may have influenced time to treatment commencement. Logistical issues and inclement weather at times affected the ability for Cairns-based TB clinicians to access the Torres Strait to review patients and commence them on treatment [22]. In contrast, the Torres and Cape TB Control Unit established in 2016, hosts an ongoing presence of TB clinicians based in the Torres Strait and allowed for a continual service where suspected case management and treatment commencement could occur at any time.

In international border settings, strength is found in collaboration between international partners and where well-defined data sharing and referral agreements are in place [36]. In collaboration with the Daru TB Program, the Torres and Cape TB Control Unit implemented new procedures to enhance recognition and streamline management of symptomatic patients [37]. As a result of improved cross-border communication, programmatic rules and strategies were implemented from 2016 to strive for safe and ethical management of shared and referred patients. These included ensuring Daru’s readiness to receive patients discharged from Australian hospitals, timely notification of microbiological and radiological results, an agreement regarding case notifications to avoid over-reporting case numbers and sharing patient outcome data. The strength of this cross-border collaboration is evidenced in this study by known treatment commencement dates of 11/12 cases from 2016.

Traditional risk factors for delays from self-reported onset of symptoms to effective treatment commencement in other settings have included limited access to health facilities, rural or remote location, and distance to diagnostics [12, 14], however in this study, these factors apply for both new and previously treated cases. With few treatment supporters to administer Directly Observed Therapy in most of the PNG Treaty villages [38] and challenging cross-border communication pathways, previous compliance history and assurances of treatment compliance to follow were not always known. It should be emphasised that none of the variables in this study were highly predictive, which shows that there is huge individual patient variation in time to effective treatment commencement and no easily identifiable risk group. It is possible that treatment delays were more prominent in patients who had previously received some treatment due to a lack of information, patient counselling and education from past TB care providers [39]. As previously treated patients may have had negative past treatment experiences thus delaying retreatment commencement, additional support for these patients is warranted [40].

To help minimise the economic burden and improve timely access for PNG patients receiving TB care in the Torres Strait between 2006 and 2012, the Cairns TB Control Unit initiated free transport for PNG Nationals accessing outreach specialist clinics on Saibai and Boigu Islands (refer to figure 1) however large numbers of patients failed to attend appointments [22]. A previous study identified that of 73 PNG MDR-TB patients diagnosed in the Torres Strait, 15 were loss to follow up, and a further 25 had an unknown treatment outcome [22]. A recent study conducted on Daru Island, reported longer delays to treatment commencement in symptomatic DR-TB household contacts presenting for TB screening when compared to symptomatic drug-susceptible household contacts [41]. This suggests that there may be country-specific risk factors which require focused support for DR-TB patients and their close contacts. It may also indicate that previously treated patients may have reduced access, a reluctance to present or have different ideologies, prompting a need for additional support for this cohort of patients.

Potential reasons for delays in previously treated cases could be that TB patients may not have been aware that they could be diagnosed twice, may have been too sick to present earlier, or may not have been experiencing any signs and symptoms at all. It is also possible that pain and adverse reactions from injectables drugs used in previous management of DR-TB acted as a deterrent for patients or their close contacts presenting earlier [42], although it should be noted that only 45.8% of MDR-TB cases diagnosed in PNG nationals in this region between 2000 and 2014 received injectables [22]. Nevertheless, as 80% of cases were new in this study, this suggests that there may be evidence of acquired drug-resistance in this population as a result of interrupted, irregular or inadequate treatment. Future qualitative research may assist TB programs in the Torres Strait to identify reasons why previously treated cases presented late in this region, and lead to improved and targeted strategies to improve early entry into the TB care pathway.

## Limitations

It is possible that extensive delays in treatment commencement were attributed to repeated patient visits to PHCs, serviced by outreach Rural Generalist Practitioners. It is possible that some symptomatic patients did not immediately enter into the TB diagnostic pathway as PHC clinicians and Rural Generalist Practitioners may not have recognised TB as a leading differential diagnosis. As a satellite TB service was provided up until 2016, it is possible that TB clinicians were not notified of all symptomatic patients and Rural Generalist Practitioners prescribed a trial of broad-spectrum antibiotics to first rule out lower respiratory tract infections [43], as has been reported in other studies [40, 44]. This study did not explore this possibility.

This study only captured PNG patients diagnosed by health services on the Australian side of the border as it focused on the management of DR-TB by the Australian Health System only. A similar study could be conducted from the PNG Health System’s perspective to capture PNG nationals diagnosed and treated by PNG health services to gain greater understanding of the overall management of DR-TB in this cross-border region. Should a study take place, a greater collaboration and linkage of data between health systems from both sides of the border would enhance the understanding of DR-TB management in the region and help improve programmatic strategies across the border.

## Conclusion

Minimising diagnostic and TB treatment delay is a global priority. Managing TB in rural and remote settings requires TB programs to strive for minimal diagnostic delay with early initiation of effective treatment. The introduction of a decentralised TB management and control structure in the Torres Strait contributed to improved service delivery and a reduction in time to effective TB treatment commencement. Our findings suggest that time to effective treatment commencement can be reduced by improved access to advanced diagnostic technology and implementation of locally-based patient-centred initiatives. Evaluating time to effective treatment as a measure of programmatic efficacy is an important and valuable public health and quality improvement approach.

## Results

## Data Availability

All data produced in the present study are not publicly available. Due to the nature of this research, restrictions apply to the availability of these data. The data custodian of Queensland Health data produced in the study is and remains the Director-General and/or the Chief Executive of the relevant Hospital and Health Service on behalf of the State of Queensland.

